# Long-term effects of directional deep brain stimulation in Parkinson’s disease: a randomized clinical trial on motor and non-motor symptoms

**DOI:** 10.1101/2023.10.30.23297793

**Authors:** A Gharabaghi, I Cebi, M Scherer, P Bookjans, B Brunnett, L Milosevic, D Weiss

**Author notes:** Corresponding author: Prof. Alireza Gharabaghi. Address: Institute for Neuromodulation and Neurotechnology, University Hospital Tübingen, Otfried-Müller-Str. 45, 72076 Tübingen, Germany.

## Abstract

**Background:** Deep brain stimulation (DBS) of the subthalamic nucleus (STN) in Parkinson’s disease (PD) that is more precisely focused to the desired target structure may avoid nearby structures that are responsible for undesired side effects.

**Objective:** Comparing the long-term effects of STN-DBS with segmented or ring contacts on motor and non-motor symptoms in akinetic-rigid PD patients.

**Methods:** This study was a prospective randomized clinical trial. At 6-months postoperatively, the optimal omnidirectional (OS) and directional (DS) stimulation contacts were compared in MedOFF within a double-blind cross-over design, both acutely (within one day) and chronically, i.e., after 3-week stimulation blocks of each condition. The examination included motor and non-motor evaluations (e.g., cognition, mood and quality of life). Importantly, the stimulation intensity of the optimal DS was adjusted such that the total electrical energy delivered (TEED) was equivalent to the TEED of the optimal OS.

**Results:** There were no significant differences between OS and DS with regard to all outcome parameters, with 30% less stimulation intensity of the latter. Notably, OS scored (non-significantly) better than DS in all motor and non-motor measures apart from the cognitive evaluation, where OS led to a deterioration of executive functions. However, in 3 of 19 patients, the stimulation intensity of DS needed to be increased above the TEED-estimated values to reach the motor benefits of OS.

**Conclusions:** Reliable comparisons between OS and DS require long-term clinical evaluations. A potential differential influence on motor and non-motor symptoms needs to be investigated in future confirmatory studies.

Registration: ClinicalTrials.gov: NCT03548506

## Introduction

Deep brain stimulation (DBS) of the subthalamic nucleus (STN) is an evidence-based and cost-effective therapy for idiopathic Parkinson’s disease (PD) patients, with superior motor, non-motor and quality of life outcomes compared to the best medical therapy once dopaminergic response fluctuations occur (1–7). The greatest therapeutic effectiveness of DBS is often achieved in the motor part of the STN and its immediate surrounding (8). However, the spread of the stimulation current into nearby structures can also lead to side effects such as muscle contractions, paresthesia, postural instability, speech impairment, reduced verbal fluency, involuntary eye movements, and impaired cognition (9–11). Among them, motor-related side effects (e.g., muscle contractions) are usually immediately detected and can often be addressed with adjusting the stimulation parameters. However, when they occur at low stimulation intensities, which are insufficient to achieve the therapeutic benefits, and when reprogramming cannot resolve the condition, subsequent surgery for lead revision may be necessary (12). In contrast, non-motor side effects (e.g., cognitive impairment) are often less apparent and remain undetected, when searching for optimal stimulation parameters in the acute setting.

Technological progress of the implanted stimulation hardware has raised new hope to overcome current limitations. Specifically, in the new generation of electrodes, the middle two of the usually four ring contacts are divided into three segments each. This makes it possible, in addition to circular/omnidirectional stimulation (OS) in the conventional ring mode, to also focus the stimulation field more precisely in the horizontal plane around one or two electrode segments with directional stimulation steering (DS). In this context, previous studies have investigated, both intraoperatively (13, 14) and extraoperatively (7, 15–17), the therapeutic windows of DS and OS, i.e., the range between the stimulation amplitude that first produces beneficial effects and the stimulation amplitude that first produces side effects. An increased therapeutic window of DS vs. OS has often been reported during these acute evaluations, thus suggesting that DS away from the structures causing side effects is in principle possible. Whether these gains are also clinically relevant in the long-term, e.g., by compensating for suboptimal electrode placements and reducing the necessity for revision surgery, is still an open question. Despite a considerable evolution of image-guided neurosurgical targeting methods (18, 19), refined intraoperative techniques (20, 21), awake procedures (22) and novel stimulation paradigms (23), the side effects and/or insufficient therapeutic benefits may lead to revision surgery in up to 15-30% of patients (12). A simulation study, for example, suggests that DS reduces out-of-target stimulation (that would lead to side effects), but is limited in achieving the target volume (for maximum therapeutic benefit), when compensating for suboptimal electrode placements with a shift of more than 1 mm (24). Moreover, a recent study revealed that six of 42 patients (i.e., ten of 72 leads) required surgical revision owing to electrode malposition despite the ability to steer the current with DS to relieve stimulation-induced side effects (25). This may suggest that the frequently described improvements in surrogate parameters such as the therapeutic window do not regularly translate into clinical benefits (26).

On this basis, it is of particular relevance to determine the potential of DS to improve the therapeutic precision in the majority of patients with well-placed electrode leads where beneficial therapeutic effects can be achieved without acute side effects. For these cases, it is unknown as to whether DS allows for a more efficient stimulation of the target structure, e.g., by lowering the stimulation intensity required to achieve the desired clinical benefit, also for longer observation periods beyond acute assessments. Such long-term assessments would allow to estimate potential benefits of DS also with regard to non-motor side effects, e.g., cognitive impairment.

In this context, an important consideration is that therapeutic impedance and surface current density are dependent on the electrode surface, and systematically differ between larger ring and smaller segmented contact sizes; associated with greater current density of the latter (27). It has therefore been suggested that stimulation intensity adjustments using DS should be performed using smaller amplitude increments (i.e., 0.1–0.3 mA compared to the traditional 0.5 mA). Moreover, it needs to be considered that because of the higher impedance (1.8-2.2 times that of standard ring contacts) due to the smaller stimulating surface of segmented contacts, stimulation at a particular intensity is associated with a greater total electrical energy delivered (TEED) when using DS compared to OS. As such, these settings would impose a greater energy burden and quicken battery consumption; which is an important factor influencing the frequency of subsequent surgical procedures for battery replacement. Already an 18% increase from the average TEED with DS vs. OS, would lead to as much as a year of battery lifespan lost ^45^. Therefore, the desired stimulation effects need to be achieved with at least the same TEED, i.e., about 30% lower stimulation amplitudes of DS vs. OS, to avoid a negative influence on the battery lifespan (28). These aspects need to be considered when aiming for overall improvements of clinical care. Moreover, the stimulation effects in previous studies were only recorded clinically using an ordinal scale (i.e., the Unified Parkinson’s Disease Ratings Scale; UPDRS) and were therefore dependent on subjective assessments of the symptoms.

We have therefore designed a study to systematically compare the efficacy of DS vs. OS in the context of a prospective, randomized, double-blind, crossover clinical trial, taking the aforementioned considerations into account. Namely, (i) applying – in addition to clinical measures - also an objective and ratio-scaled measure, i.e., electromyography (EMG) (29) to assess muscular rigidity; (ii) conducting stimulation titration at each contact with small increments in stimulation intensity (0.2 mA steps) for both stimulation conditions; (iii) maintaining TEED between DS and OS; (iv) applying the randomized, double-blinded evaluations after both acute *and* chronic stimulation periods; and (v) assessing both motor *and* non-motor symptoms. We tested the hypothesis that, even under these rigorously controlled conditions, the focused stimulation of the target structure with DS could achieve similar therapeutic efficacy on motor symptoms while reducing non-motor side effects.

## Methods

### Patients

All patients who participated in this study were nominated for deep brain stimulation surgery upon multidisciplinary review based on standard inclusion/exclusion criteria for STN-DBS in PD (30), independent from study participation. The study was approved by the ethics committee of the Medical Faculty of the University of Tuebingen. Twenty-three consecutive akinetic-rigid patients scheduled for STN-DBS were included in this study after providing written informed consent to participation. Four patients dropped out due to personal reasons not related to the study. Nine-teen patients completed the study with the cross-over treatment evaluation periods. Participant and drop-out numbers at each phase of the study are summarized in Figure. 1. Patient demographics are summarized in Table 1.

**Figure 1:**
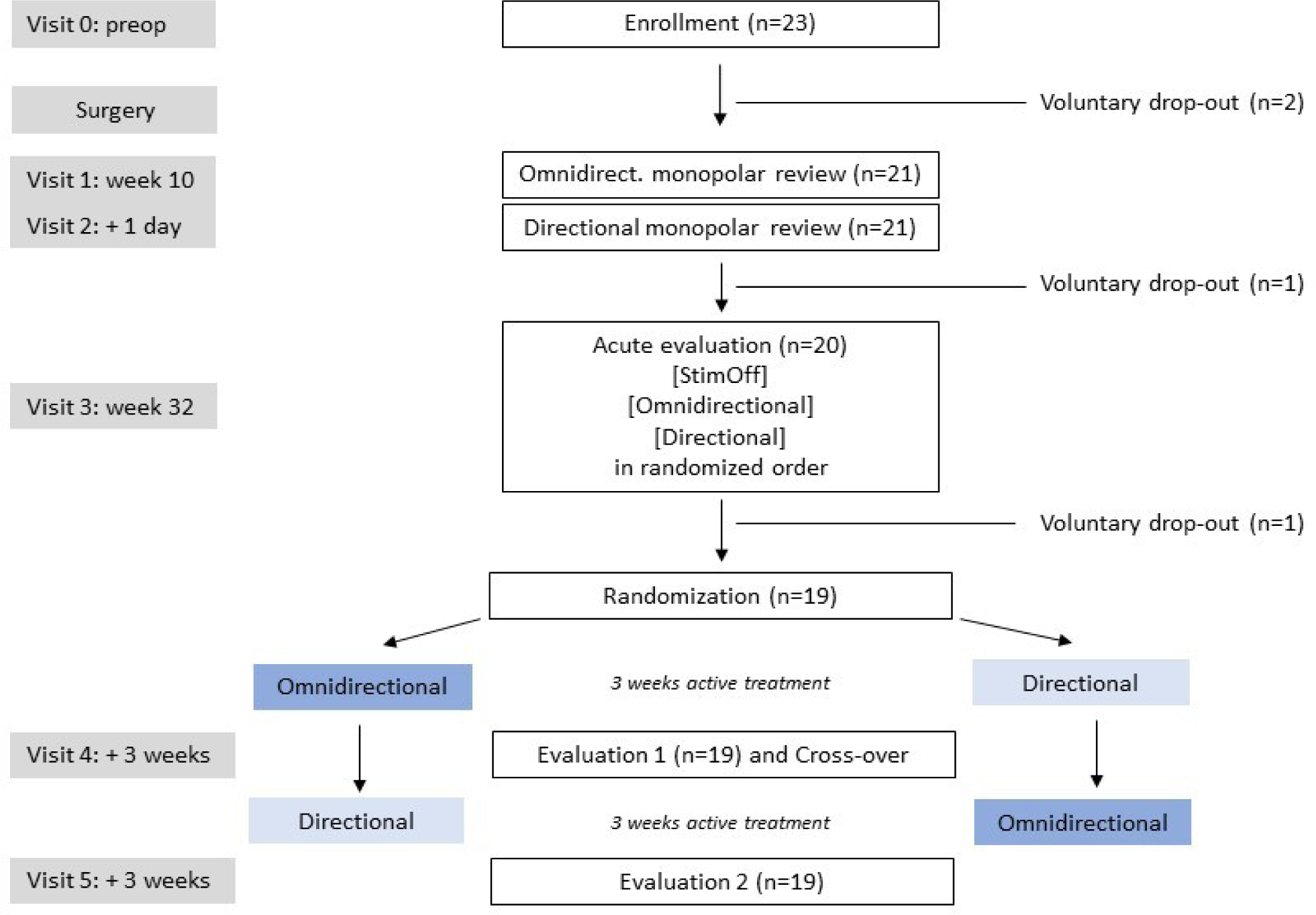
Consort flow diagram of the SANTOP study

**Table 1:**
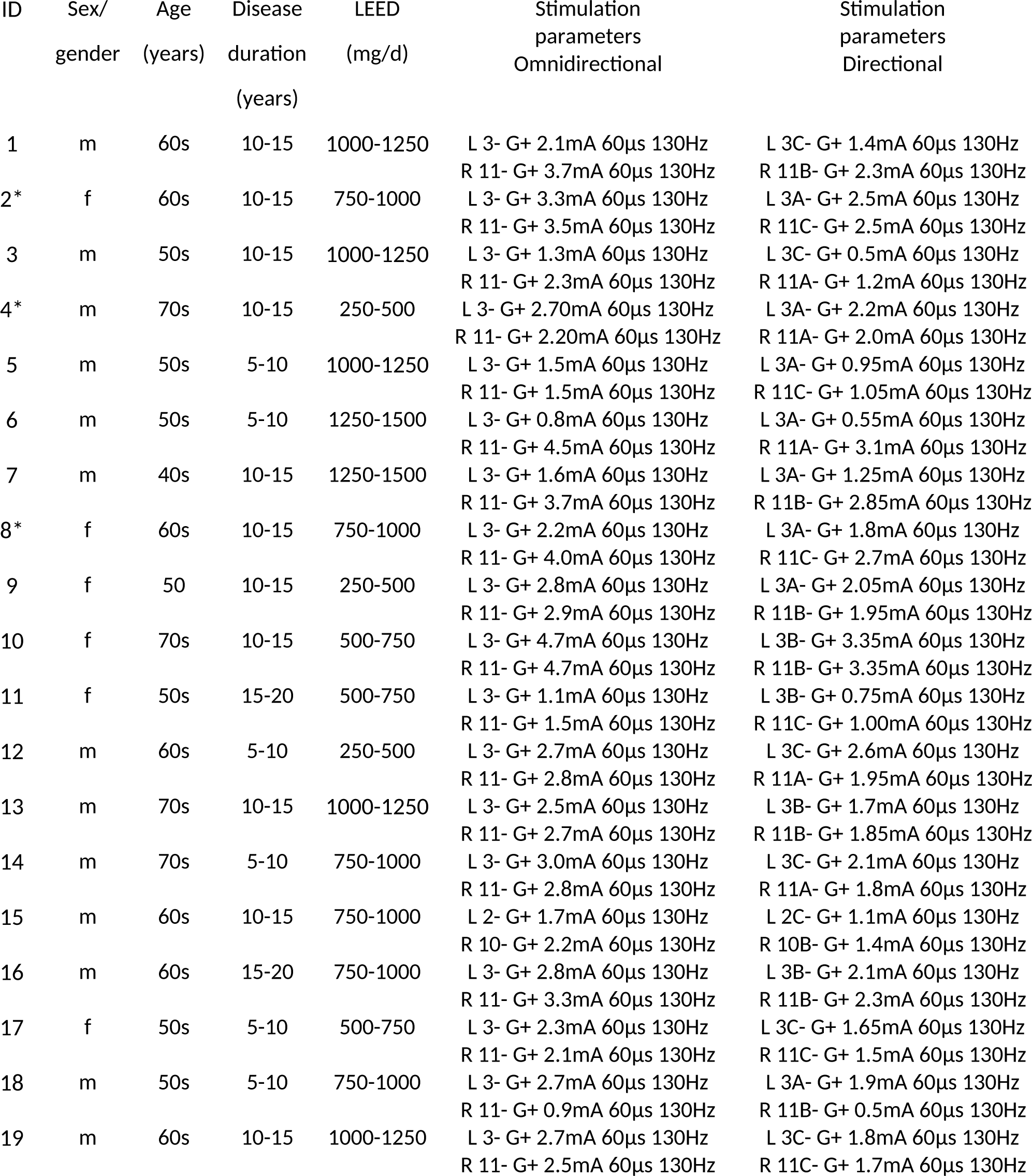
Patient demographics and stimulation parameters are summarized in. LEED indicates the levodopa equivalent dose during the cross-over-phase. The asterisk “*” indicates patients in whom a reprogramming was necessary during the cross-over-phase: ID2: the calculated directional stimulation was L 3A- G+ 2.1mA 60µs 130Hz, R 11C- G+ 2.2mA 60µs 130Hz; patient reported bradykinesia and had rigidity and freezing of gait with directional stimulation during the cross-over-phase so that we needed to increase the stimulation Intensity. ID4: the calculated directional stimulation was L 3A- G+ 1.85mA 60µs 130Hz, R 11A- G+ 1.55mA 60µs 130Hz; patient reported bradykinesia and had rigidity with directional stimulation during the cross-over-phase so that we needed to increase the stimulation intensity. ID8: the calculated directional stimulation was L 3A- G+ 1.6mA 60µs 130Hz, R 11C- G+ 2.6mA 60µs 130Hz; patient reported worsening of motor fluctuations so that we needed to increase the stimulation intensity.

### Surgical procedure

The tentative implantation location in each hemisphere was initially determined by conventional image-based direct targeting of the STN using preoperative MRI (T2-weighted and/or SWI) (31). Patients were withdrawn from antiparkinsonian medications overnight, and on the day of surgery electrophysiological recordings of single-unit and/or local field potential activities were used to confirm the target location in each hemisphere. Initially, single unit recordings were used to delineate STN entry and spatial extent by characteristic burst firing rates (20-60 Hz) and irregular or bursting patterns (32). The trajectory yielding greatest spatial extent of STN was then selected for the DBS electrode implant. The final position of the DBS electrode was once again titrated using neurophysiological recordings directly from the DBS contacts, such that two levels of the DBS lead containing segmented contacts were placed in the area of STN yielding greatest beta (13-30 Hz) oscillatory power, determined by (online) intraoperative power spectral density calculations (21). There were no complications from surgical procedures.

### Study design

This study was a prospective, randomized, double-blind, crossover, single-center clinical trial, i.e., the SANTOP (Subthalamic steering for therapy optimization in Parkinson’s Disease) study (preregistered at ClinicalTrials.gov: NCT03548506). The comprehensive study design is illustrated as Figure 1. The patients were enrolled preoperatively (V0). The postoperative visits started 2-months (V1/2) and 6-months (V3/4/5) after surgery. All evaluations were done in the Med OFF condition after overnight withdrawal of dopaminergic medication. The patients and the evaluators were blinded to stimulation conditions during the whole study. At the 2-months visits, a monopolar review was performed on two subsequent days to identify the optimal contact and stimulation parameters for omnidirectional (V1) and directional (V2) stimulation.

At the 6-months visit (V3), OS and DS were compared to each other and to Stim OFF in randomized order, i.e., 30 minutes from reprogramming for each condition (acute setting). Afterwards, stimulation settings were randomized again for a three-week (chronic setting) active treatment period (by a study coordinator who would not be involved in clinical evaluation) to either the omnidirectional settings, or the energy-equivalent directional settings (preconfigured options in the stimulation programmer). After the first three-week treatment period (V4), blinded collection of outcome data (described below) was performed by a clinician, followed by a cross-over in stimulation settings performed by a study coordinator. After another 3-week treatment period (V5), blinded outcome data were collected once more. This procedure allowed to directly compare OS and DS after long-term stimulation periods, during which other parameters (such as medication) were kept unchanged. These study epochs are summarized in Figure 1.

### Identification of optimal contact and stimulation parameters

At 2-months (on average 67 days, +/- 9 days, range 56-88 days) postoperatively (see Figure 1), stimulation titration was performed on two subsequent days after overnight withdrawal from medication. On the first day (V1), each patient underwent a monopolar review of the three upper contacts using circular stimulation at each level to determine the optimal OS level. On the second day (V2), the same procedure was repeated using the three segmented contacts of the previously determined best omnidirectional level (V1) to determine the optimal stimulation direction.

During these visits, rigidity in the contralateral upper limb was assessed (double-blind) by a clinician during unilateral stimulation, while the programming and random electrode contact selection was conducted by another examiner at a stimulation frequency of 130 Hz and a pulse width 60 μs, using intensities starting from 0.5 mA and incrementally increasing by 0.2 mA, until a (self-reported) side effect threshold was reached. At each stimulation increment, a 1-minute rigidity assessment (consisting of passive movements of the upper limb by flexing and extending the elbow) was performed to determine the effect threshold.

The therapeutic window was determined for each evaluated contact, and the contact with the greatest therapeutic window and/or best effect threshold in each hemisphere was considered optimal. The stimulation intensity for each optimal segmented contact was then determined by titration of the stimulation intensity such that the total electrical energy delivered (33) was equivalent to the total electrical energy of the optimal omnidirectional settings. Of note, the stimulation intensity for DS was always lower than for OS due to the greater impedance associated with a smaller stimulation surface (see also the Introduction). The OS and DS parameters for each patient are available in Table 1.

### Treatment evaluation and outcome measures

At a minimum of 6-months (on average 229 days, +/- 46 days, range 187-369 days) postoperatively (see Figure 1), OS and DS were compared to each other in an acute (V3) and chronic setting (V4/5). The muscle rigidity of the upper extremity was measured by means of surface EMG recording (BrainAmp DC/EXG, Brain Products, Munich, Germany; sampled at 5 kHz) of the M. biceps brachii and the M. triceps brachii. Previous work suggests that the EMG profile during standardized passive extension and flexion movements of the elbow joint is influenced by DBS and corresponds with standardized clinical scales (29). The evaluation procedure consisted of EMG recording during a 30-second non-movement period, followed by a 60-second period of passive flexion/extension (∼0.5 Hz cycles) of the elbow (∼90°). Five trials of this procedure were performed for each arm at each stimulation intensity and each electrode contact.

At both V3 and V4/5, the following data was collected: MDS-UPDRS parts III (34); timed Capsit-PD walking test (35); Freezing of Gait Assessment Course (FoG-AC) (36); Spatial (SDR) (37) and verbal (VDR) (38) working memory with respective visual (39) and acoustic odd-ball tests (40) for attention monitoring, and task-related EEG recordings (41). During the acute V3 assessment, no stimulation (STIM OFF), directional and omnidirectional settings were compared in randomized order. During the chronic V4/5 cross-over assessment, directional and omnidirectional settings were compared in randomized order.

At V4/5, this additional data was acquired: MDS-UPDRS part I, II and IV (34); Parkinson’s Disease Quality of Life Scale (PDQ-39) (42); Becks ‘Depression Inventory (BDI) (43); Montreal Cognitive Assessment (MoCA) (44); Apathy Scale (45); Lille Apathy rating scale (46); Deep Brain Stimulation impairment scale (DBS-IS) (47).

### EMG data processing and analysis

EMG data processing consisted of artifact rejection, epoching, feature extraction and, faulty trial rejection. Initially, the stimulation artifact, the line noise, and their respective harmonics were removed from the data. After artifacts had been removed, the data were split into epochs. From each run, two segments were extracted, a 30-second non-movement segment and a 30-second movement segment. These segments were further divided into individual 2-second epochs. While strict 2-second cutting was applied to the non-movement phase data, the movement phase data were cut according to full movement cycles (0.5 Hz). To extract feature information from the EMG signal, the signal was detrended, and enveloped via the absolute of the Hilbert transform. The average amplitude (power) of the signal was extracted as feature information and was then normalized with respect to the overall EMG power between 2 and 300 Hz to account for different impedance levels across recordings. Finally, outliers on a feature level were rejected if z-score > 2 (i.e., more than two standard deviations from the mean).

EMG data was analyzed using generalized linear mixed models (glmm). Factors modeled in the analysis were as follows: “counter-movement” to evaluate the effect of the counter-movement on the EMG power; the “stimulation-condition:” to contrast DS and OS modes; “movement” to contrast between rest versus passive movement phases in EMG activity; the interaction between the stimulation-condition and movement; the (chronologically ordered) “recording id” to compensate for repeated measurements over time; “trial id” to compensate for repeated measurements within trials; “patient id” as a random factor to compensate for repeated measurements across patients. Secondary outcome data of standardized clinical score assessments were assessed using paired Wilcoxon signed-rank tests to compare stimulation conditions.

## Results

At 6-months postoperatively, optimal ring and segmented contacts were compared to each other with average TEED-adapted stimulation intensities of 2.59 ± 0.77 mA for OS and 1.82 ± 0.61 mA for DS; this was equivalent to a difference of 30.37 ± 8.47 %. First, OS and DS were compared to each other and to Stim OFF in randomized, cross-over order in an acute setting (V3), i.e., 30 min after reprogramming within the same day. After this acute assessment, OS and DS were compared to each other in randomized, cross-over order in a chronic/long-term setting, i.e., 3 weeks after reprogramming for each condition (V4/5).

### Motor symptoms

In the *acute setting* (V3), both conditions had similar therapeutic efficacy on motor symptoms, i.e., there was no significant difference between OS and DS (Table 2). Specifically, OS (30.00 ± 13.55; p=.001) and DS (30.72 ± 10.87; p=.003) improved UPDRS III as compared to Stim OFF (39.63 ± 12.92); also, the FoG-AC improved with both OS (14.83 ± 13.66; p=.021) and DS (13.92 ±13.32; p=.024) as compared to Stim OFF (17.83 ± 15.05). There were, however, no significant differences between OS and DS for UPDRS III (p=.521) and FoG-AC (p=.591), or the other outcome measures including the oddball and working memory tasks (VDR/SDR).

**Table 2:**
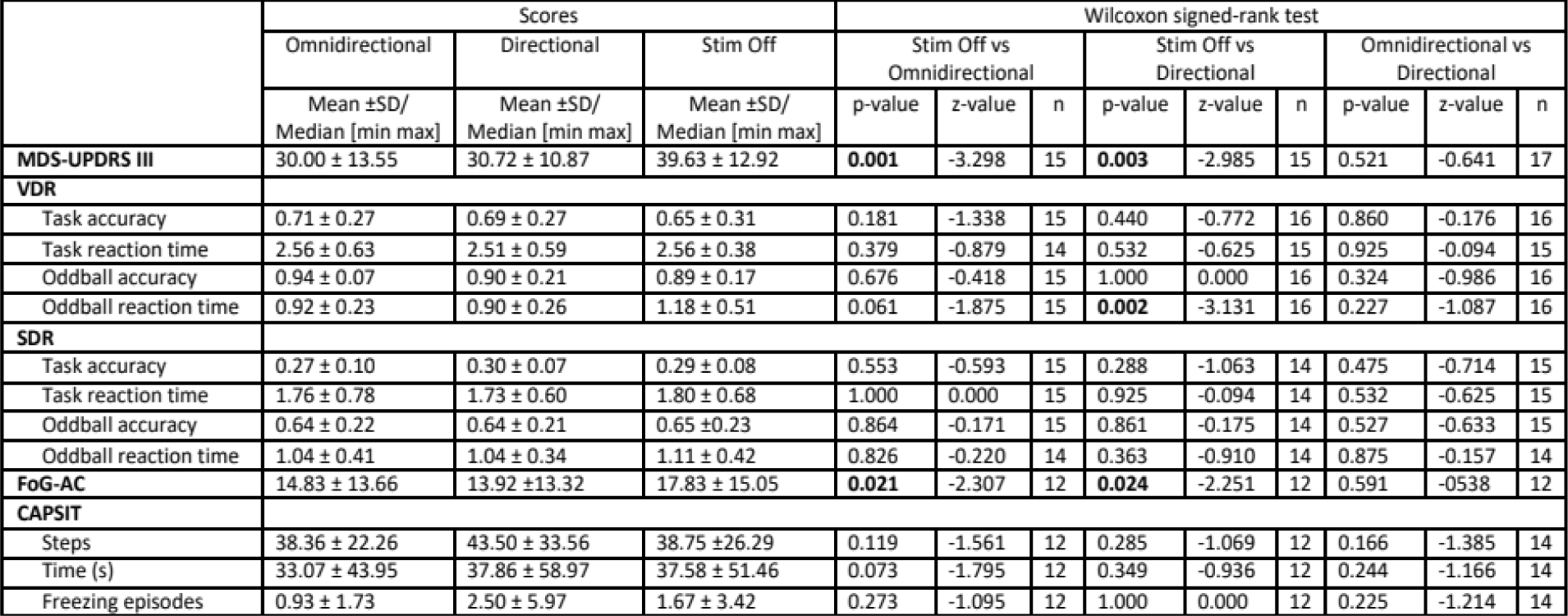
Six months after surgery (V3), both conditions (OS and DS) had significant therapeutic efficacy with regard to segmental (MDS-UPDRS III) and axial motor (FOG-AC) symptoms compared to Stim Off during the acute evaluation (i.e., 30 min stimulation periods); however, there were no significant differences between the omnidirectional (OS) and directional (DS) conditions.

In the *long-term setting* (V4/5), both conditions had also similar therapeutic efficacy, i.e., there were no significant differences between OS and DS with regard to motor symptoms (Table 3) and EMG-based assessments (p ≥ .135). However, in 3 of the 19 patients (i.e., 15.79%), stimulation amplitudes had to be increased for the DS condition (above and beyond the estimated TEED) to achieve the therapeutic current strength, i.e., to avoid motor fluctuations, bradykinesia, rigidity and freezing of gait (for further details see the legend of Table 1).

**Table 3:** Six months after surgery, in the cross-over period (V4/5), there were no significant differences between OS and DS with regard to motor and non-motor symptoms during the chronic evaluation (i.e., after 3-week stimulation blocks of each condition).

|  | Scores |  | Wilcoxon signed-rank test |  |  |
| --- | --- | --- | --- | --- | --- |
|  | Omnidirectional | Directional | Omnidirectional vs Directional |  |  |
| | Mean ±SD/<br>Median [min max] | Mean ±SD/<br>Median [min max] | p-value | z-value | n |
| **MDS-UPDRS I** | 11.16 ± 8.49 | 11.00 ± 5.94 | 0.909 | -0.114 | 17 |
| **MDS-UPDRS II** | 18.16 ± 9.32 | 19.06 ± 9.19 | 0.568 | -0.571 | 18 |
| **MDS-UPDRS III** | 28.63 ± 12.91 | 32.42 ± 15.07 | 0.121 | -1.550 | 19 |
| **MDS-UPDRS IV** | 8.17 ± 4.89 | 8.78 ± 4.45 | 0.925 | -0.095 | 17 |
| **VDR** |  |  |  |  |  |
| Task accuracy | 0.70 ± 0.29 | 0.69 ± 0.23 | 1.000 | 0.000 | 17 |
| Task reaction time | 2.56 ± 0.51 | 2.54 ± 0.59 | 0.845 | -0.196 | 18 |
| Oddball accuracy | 0.96 ± 0.08 | 0.94 ± 0.11 | 0.271 | -1.100 | 18 |
| Oddball reaction time | 0.91 ± 0.32 | 0.85 ± 0.27 | 0.267 | -1.111 | 18 |
| **SDR** |  |  |  |  |  |
| Task accuracy | 0.25 ± 0.12 | 0.22 ± 0.09 | 0.355 | -0.924 | 17 |
| Task reaction time | 1.83 ± 0.64 | 1.71 ± 0.53 | 0.918 | -0.103 | 16 |
| Oddball accuracy | 0.64 ± 0.26 | 0.57 ± 0.26 | 0.265 | -1.115 | 17 |
| Oddball reaction time | 1.09 ± 0.35 | 1.01 ± 0.33 | 0.469 | -0.724 | 16 |
| **FoG-AC** | 11.47 ± 13.02 | 12.80 ± 13.80 | 0.129 | -1.517 | 15 |
| **CAPSiT** |  |  |  |  |  |
| Steps | 39.11 ± 20.55 | 42.00 ± 22.02 | 0.072 | -1.801 | 17 |
| Time (s) | 27.44 ± 23.74 | 31.82 ± 34.04 | 0.142 | -1.468 | 17 |
| Freezing episodes | 1.56 ± 3.75 | 1.53 ± 3.63 | 0.131 | -1.511 | 17 |
| **MOCA** | 26.53 ± 3.15 | 27.83 ± 1.92 | 0.080 | -1.753 | 17 |
| Executive | 3.89 ± 1.05 | 4.47 ± 0.84 | 0.071 | -1.805 | 19 |
| Naming | 2.84 ± 0.69 | 3.00 ± 0.00 | 0.317 | -1.000 | 18 |
| Attention | 5.58 ± 0.69 | 5.44 ± 0.86 | 0.206 | -1.265 | 18 |
| Language | 2.44 ± 0.62 | 2.32 ± 0.82 | 0.366 | -0.905 | 18 |
| Abstraction | 1.89 ± 0.32 | 2.00 ± 0.00 | 0.157 | -1.414 | 18 |
| Delayed Recall | 3.95 ± 1.31 | 4.37 ± 0.83 | 0.323 | -0.988 | 19 |
| Orientation | 5.95 ± 0.23 | 6.00 ± 0.00 | 0.317 | -1.000 | 19 |
| **BDI** | 13.56 ± 7.25 | 14.88 ± 6.91 | 0.448 | -0.758 | 17 |
| **AS** | 16.11 ± 4.58 | 17.58 ± 4.49 | 0.646 | -0.406 | 16 |
| **DBS-IS** | 25.94 ± 14.61 | 28.06 ± 12.52 | 0.674 | -0.421 | 17 |
| **LARS** | -22.47 ± 7.33 | -20.61 ± 10.14 | 0.275 | -1.091 | 18 |
| **PDQ 39** |  |  |  |  |  |
| Mobility | 19.22 ± 10.73 | 21.13 ± 8.16 | 0.380 | -0.877 | 15 |
| Activities of daily living | 9.06 ± 5.68 | 9.72 ± 5.06 | 1.000 | 0.000 | 17 |
| Emotions | 8.06 ± 4.56 | 9.00 ± 4.04 | 0.318 | -0.999 | 17 |
| Stigma | 3.50 ± 3.24 | 4.39 ± 2.43 | 0.304 | -1.027 | 17 |
| Social support | 1.94 ± 2.31 | 2.53 ± 2.27 | 0.094 | -1.672 | 17 |
| Cognition | 5.00 ± 3.27 | 6.38 ± 2.90 | 0.477 | -0.711 | 15 |
| Communication | 4.06 ± 3.42 | 4.53 ± 2.74 | 0.832 | -0.212 | 16 |
| Bodily discomfort | 5.11 ± 2.61 | 5.35 ± 2.34 | 0.640 | -0.467 | 16 |

Notably, OS scored (non-significantly) better than DS for UPDRS III (28.63 ± 12.91 vs. 32.42 ± 15.07), FoG (11.47 ± 13.02 vs. 12.80 ± 13.80), CAPSIT steps (39.11 ± 20.55 vs. 42.00 ± 22.02), CAPSIT time (27.44 ± 23.74 vs. 31.82 ± 34.04), and PDQ-39 Mobility (19.22 ± 10.73 vs. 21.13 ± 8.16). Moreover, long-term stimulation (i.e., evaluation after 3-week blocks, V4/5) changed the scores in comparison to the acute evaluation (i.e., evaluation after 30 min, V3); specifically, the UPDRS III score improved from 30.00 ± 13.55 to 28.63 ± 12.91 for OS, and deteriorated from 30.72 ± 10.87 to 32.42 ± 15.07 for DS.

### Non-motor symptoms

In the long-term setting (V4/5), there were no significant differences between OS and DS with regard to non-motor symptoms (Table 3). Notably, OS scored (non-significantly) better than DS on all (BDI, AS, DBS-IS, LARS, PDQ-39) but one non-motor clinical measure. The only non-motor clinical measure favoring DS was the MOCA evaluation; specifically, OS led to (non-significantly) more cognitive impairment, i.e., a lower MOCA score as compared to DS (26.53 ± 3.15 vs. 27.83 ± 1.92; p=.080). This effect was driven by a deterioration of the MOCA sub-score for executive functions (OS vs. DS: 3.89 ± 1.05 vs. 4.47 ± 0.84; p=.071; OS vs. preoperative baseline: 3.89 ± 1.05 vs. 4.63 ± 0.60; p=.015).

## Discussion

This study determined the potential of directional stimulation steering (DS) with segmented contacts to improve the therapeutic precision of STN-DBS. Specifically, and in contrast to previous work, we investigated whether DS allowed for a more efficient stimulation of the target structure by lowering the stimulation intensity required to achieve the clinical benefit with regard to motor symptoms. For this purpose, the study had unique methodological features: The optimal contacts were identified by conducting the stimulation titration with small increments in stimulation intensity (0.2 mA steps) for both OS and DS; the TEED was maintained between OS and DS for longer observation periods beyond acute assessments in a cross-over design; thereby, also non-motor symptoms, e.g., cognitive impairment, could be assessed. By that, this study revealed that there were no significant differences between OS and DS with regard to all outcome parameters, with 30% less stimulation intensity of the latter. Whether the more focused stimulation of the target structure with DS will also achieve fewer side effects, e.g., with regard to executive functions, needs to be investigated in confirmatory studies, since this exploratory approach could potentially be affected by multiple comparisons.

Currently, there are few systematic studies comparing directional STN-DBS to standard omnidirectional stimulation which include intra- and extra-operative assessments. One intraoperative study (14) demonstrated a larger therapeutic window with DS, i.e., an increase of the distance between the clinical effect threshold (i.e., minimum stimulation intensity to elicit a beneficial effect) and the side effect threshold (i.e., minimum stimulation intensity to elicit side effects). However, another intraoperative study (13) did not demonstrate any relevant increases of the therapeutic window with DS compared to conventional OS. Two extraoperative studies (one retrospective, open-label pilot study (16), and one prospective, randomized and double-blind study (15)) found an increase in the side effect threshold, but no decrease in the clinical effect threshold (i.e., therapeutic current strength); while another extraoperative retrospective, open-label case series showed a decrease also of the clinical effect threshold with directional programming (48). However, the clinical evaluations in this latter study were conducted with different step-sizes of the applied stimulation amplitudes, i.e., 0.5 mA for OS and 0.1-0.2 mA for DS, which considers the higher charge density of the latter, but may also bias the findings with regard to the sensitivity for clinical effect and side effect threshold detection. An accurate thresholding with the same amplitude step-sizes for both stimulation conditions is an important prerequisite for detecting improved therapeutic effectiveness of one approach or the other. Furthermore, relevant differences between the various segmented stimulation directions could be found with regards to the side effect threshold (16), but not with regard to the therapeutic effectiveness (14). Possible reasons for these variable findings are: (i) different accuracies in titration of the stimulation amplitude (in previous extraoperative studies, the systematic titration of the stimulation amplitude was carried out in steps of 0.5 mA (16) or 1.0 mA (15), whereas Pollo and colleagues (14) chose 0.1 mA steps for their intraoperative examinations); (ii) stimulation effects were recorded clinically and on an ordinal scale (i.e. UPDRSIII) and were therefore dependent on a subjective assessment of the symptoms and the experience of the examiner.

The most comprehensive investigation to date is the prospective, randomized, double-blind PROGRESS multicenter study (17). The primary endpoint was evaluated acutely three months after initial programming by demonstrating that DS resulted in a wider therapeutic window using DS (2.98 ± 1.38 mA) compared to OS (2.11 ± 1.33 mA). Moreover, DS (1.11 ± 1.00 mA) could reduce the current required to achieve symptom relief compared to OS (1.83 ± 1.52 mA) (17). As with previous studies, a systematic evaluation of non-motor symptoms was missing, and unlike our study, the outcome was based on subjective clinical assessments using the ordinal UPDRS scale only. Also, the stimulation titration protocols were not reported in the study, and presumably varied across centers. And finally, like all studies to date that have systematically assessed directional STN-DBS, the TEED across stimulation conditions was not maintained for longer periods. Moreover, the 3-months open-label observations had a fixed order (i.e., OS period always prior to the DS period).

The present study aimed to address some limitations of previous studies investigating directional DBS. Namely, OS and DS were compared by complementing the subjective clinical evaluation by an objective assessment of muscular rigidity using EMG; furthermore, stimulation parameter titration was performed using small increments in the current amplitudes for both stimulation conditions; and finally, TEED, and thereby battery power consumption, was maintained for three weeks for each condition, when comparing OS and DS.

In the present study, DS and OS had no significant differences with regard to motor symptoms in both clinical and EMG-based measures, when controlling for the above-mentioned confounding factors. However, OS had (non-significantly) better clinical effects than DS on all motor and non-motor measures apart from the cognitive evaluation (see below). These differences between OS and DS were more apparent in the chronic (3 weeks) than in the acute (30 minutes) evaluation settings.

Specifically, the UPDRS III score (39.63 ± 12.92 in the Stim OFF condition) improved with OS (30.00 ± 13.55; p=.001) and DS (30.72 ± 10.87; p=.003) in the acute assessment. In the chronic setting, UPDRS III improved even further with OS (28.63 ± 12.91), while it deteriorated with DS (32.42± 15.07); notably, the average difference (28.63 ± 12.91 vs. 32.42± 15.07) between conditions crossed the 3.25-point threshold for a minimal clinically important difference (MCID) of the UPDRS III (49), even though this difference was not significant due to the large variability within each condition.

Similarly, FoG-AC (17.83 ± 15.05 in the Stim OFF condition) improved with both OS (14.83 ± 13.66; p=.021) and DS (13.92 ±13.32; p=.024) in the acute assessment, and even further in the chronic setting (OS: 11.47 ± 13.02; DS: 12.80 ± 13.80).

These observations were complemented by the fact that in 3 of 19 patients, the electrical energy of DS needed to be increased during the chronic/long-term period above and beyond the estimated TEED to reach the therapeutic motor benefits of OS. These findings indicate that long-term assessments are necessary to capture the overall benefit of different programming approaches.

The cognitive long-term assessment revealed an exception and the only difference between the stimulation conditions favoring DC; namely, DS led to (non-significantly) higher MOCA scores, i.e., less cognitive impairment, as compared to OS (27.83 ± 1.92 vs. 26.53 ± 3.15; p=.080). Despite the small effect size, this observation warrants further investigations in future due to different considerations: This finding was the only one in favour of DS, and thereby in contrast to the above-mentioned overall trend of the other motor and non-motor findings in favour of OS. Moreover, this MOCA change in PD patients is not to be expected within the short six weeks evaluation period (50). Furthermore, the observed effect was driven by a deterioration of the MOCA sub-score for executive function (OS vs. DS: 3.89 ± 1.05 vs. 4.47 ± 0.84; p=.071; OS vs. preoperative baseline: 3.89 ± 1.05 vs. 4.63 ± 0.60; p=.015), which was previously shown to be particularly affected by STN-DBS (51).

This stimulation-induced impact on executive function may be explained by the different volumes of tissue activated (VTA) by OS vs. DS. Specifically, computational modeling suggested that TEED-balanced DS, i.e., applying about 30% less stimulation amplitude with DS than OS (like in the present study), reduced the total VTA by 15% (28). This reduced the coverage of the targeted and non-targeted volumes in the STN leading to less motor benefits and less side effects regarding executive function, respectively. However, these interpretations remain speculative since the clinical differences between the OS and DS conditions did not reach statistical significance.

### Limitations and considerations

While we attempted to control as many conditions as possible to achieve an impartial and highly systematic comparison between DS and OS, this study is not without limitations. It must first be acknowledged that the sample size of our study is comparable to other single center studies but far smaller than that of the multicenter PROGRESS study. However, one advantage of our study is that each patient underwent a consistent and highly scrutinous procedure for stimulation programing and parameter titration using small current intensity increments for both stimulation conditions.

Moreover, the EMG activity recorded in our study provided an objective measure, but may have missed differences between conditions due to much variability between data points caused by some variability of electrode placements across evaluations. Nevertheless, the results are congruent with the standard subjective ordinal scale-based evaluations in this study, e.g., the UPDRS III scores.

Finally, a very important consideration that is unique to this study is that the TEED was maintained across the two stimulation conditions. This means that the current intensity used for DS was always lower than for OS, and lower than its own side effect threshold. While this did allow for an impartial comparison of the two conditions when taking into consideration TEED and thus battery lifespan, it did not allow for the comparison of the two conditions at each of their respective maximum current intensities, i.e., activated VTA. The current findings, therefore, suggest that DS even below the maximum tolerable settings can achieve similar therapeutic efficacy as OS, even though the OS effects seemed (non-significantly) larger across measures. However, we are left with the question of whether DS at the maximum possible current intensity may have produced superior clinical benefits in our cohort compared to OS. Finally, the observation periods of 3 weeks for each condition might have been too short to delineate even larger differences between the conditions.

### Conclusions

There were no significant differences between OS and DS with regard to motor and non-motor outcome parameters when they were applied at the same TEED levels, i.e., with 30% less stimulation intensity of the latter. However, in some cases, the stimulation intensity of DS needed to be increased above and beyond the TEED-estimated value to reach the motor benefits of OS. Furthermore, OS scored (non-significantly) better than DS in all motor and non-motor measures apart from the cognitive evaluation, where OS led to a deterioration of executive functions.

Long-term observation periods beyond acute assessments are necessary to identify differences in overall treatment benefit. Future confirmatory studies with larger sample cohorts and longer follow-up periods need to investigate the potential outcome differences of OS and DS with regard to motor and cognitive functions.

## Data Availability

All data produced in the present study are available upon reasonable request to the corresponding author

## Credit authorship contribution statement

Alireza Gharabaghi: Funding acquisition, Conceptualization, Project administration, Writing – original draft.

Idil Cebi: Project administration, Investigation, Formal analysis, Writing – review & editing.

Maximilian Scherer: Methodology, Investigation, Formal analysis, Writing – review & editing.

Patrick Bookjans: Investigation, Formal analysis, Writing – review & editing.

Bastian Brunnett: Investigation, Formal analysis, Writing – review & editing.

Luka Milosevic: Investigation, Formal analysis, Methodology, Writing – original draft.

Daniel Weiss: Funding acquisition, Conceptualization, Writing – review & editing.

## Acknowledgements

A.G. was supported by the German Federal Ministry of Education and Research (BMBF). M.S. was supported by the Alexander von Humboldt Foundation. D.W. was supported by the German Research Council (WE5375/1-3) and the Michael J Fox Foundation. We acknowledge support by the Open Access Publishing Fund of the University of Tübingen.

